# Microbial determinants of dementia risk in subjects of Mexican descent with type 2 diabetes living in South Texas

**DOI:** 10.1101/2024.03.20.24304637

**Authors:** Lisa M. Matz, Nisarg S. Shah, Laura Porterfield, Olivia M. Stuyck, Michael D. Jochum, Rakez Kayed, Giulio Taglialatela, Randall J. Urban, Shelly A. Buffington

## Abstract

Type 2 diabetes (T2D) is a common forerunner of neurodegeneration and dementia, including Alzheimer’s Disease (AD), yet the underlying mechanisms remain unresolved. Individuals of Mexican descent living in South Texas have increased prevalence of comorbid T2D and early onset AD, despite low incidence of the predisposing APOE-ε4 variant and an absence of the phenotype among relatives residing in Mexico – suggesting a role for environmental factors in coincident T2D and AD susceptibility. Here, in a small clinical trial, we show dysbiosis of the human gut microbiome could contribute to neuroinflammation and risk for AD in this population. Divergent Gastrointestinal Symptom Rating Scale (GSRS) responses, despite no differences in expressed dietary preferences, provided the first evidence for altered gut microbial ecology among T2D subjects (sT2D) *versus* population-matched healthy controls (HC). Metataxonomic 16S rRNA sequencing of participant stool revealed a decrease in alpha diversity of sT2D *versus* HC gut communities and identified BMI as a driver of gut community structure. Linear discriminant analysis effect size (LEfSe) identified a significant decrease in the relative abundance of the short-chain fatty acid-producing taxa *Lachnospiraceae*, *Faecalibacterium*, and *Alistipes* and an increase in pathobionts *Escherichia-Shigella*, *Enterobacter*, and *Clostridia innocuum* among sT2D gut microbiota, as well as differentially abundant gene and metabolic pathways. These results suggest characterization of the gut microbiome of individuals with T2D could identify key actors among “disease state” microbiota which may increase risk for or accelerate the onset of neurodegeneration. Furthermore, they identify candidate microbiome-targeted approaches for prevention and treatment of neuroinflammation in AD.

**Importance:** Mexican Americans are at increased risk for developing type 2 diabetes (T2D) that precedes Alzheimer’s Disease (AD), compared to non-Hispanic whites; however, the reason remains unknown. The leading risk factor for T2D is obesity. Among Texans, individuals of Mexican descent are disproportionately affected by obesity and T2D. Mexican immigrants to the US and their descendants face significant environmental pressures, including dietary changes. Diet is the primary determinant of gut microbiome composition, which is increasingly linked to both metabolic and brain health. Here, we performed a case-control, cross-sectional observational clinical study to test the hypothesis that diet-driven shifts in gut microbiome composition contribute to T2D and AD susceptibility in individuals of Mexican descent living in South Texas. Our results identify a microbial signature, characterized by decreased short-chain fatty acid producers with an increase in opportunistic pathogenic species, that could contribute to the increased risk for neurodegenerative disorders among individuals with T2D.

## Introduction

Epidemiological studies identify type 2 diabetes (T2D) as a common antecedent of pathological neurodegeneration and accompanying dementia, including Alzheimer’s Disease (AD)^1–3^; however, the mechanisms by which T2D increases risk for neurodegenerative disorders remains unknown. To investigate the causal relationship between T2D and comorbid AD susceptibility, we performed a small clinical trial in a population disproportionately affected by the disorders, individuals of Mexican descent living in the United States. While it is well established that this population is at increased risk of developing both T2D and, subsequently, comorbid AD compared to non-Hispanic whites^4, 5^, the contributing factors and underlying pathophysiology remain unknown. The leading risk factor for T2D is obesity^6, 7^. According to the Centers for Disease Control^8^, Texas ranks 14^th^ in the nation for obesity prevalence^9^. Within the Texan population, individuals of Mexican descent are disproportionately afflicted by obesity and T2D, with twice the prevalence of T2D (15.7%) relative to non-Hispanic whites^10–12^. Although a combination of genetic and environmental factors contribute to the etiology and pathophysiology of obesity^7^ and comorbid T2D^7, 13, 14^, a recent study of 132 twin pairs found that – independent of genetics – overnutrition is the main factor underlying higher body mass index (BMI)^15^. Furthermore, despite similar genetics, the <u>H</u>ealth and <u>A</u>ging <u>B</u>rain among <u>L</u>atino <u>E</u>lders (HABLE) study found the prevalence of abdominal obesity and T2D in Americans of Mexican descent to be significantly higher than in similarly aged patients in a strictly Mexican cohort in the Mexican Health and Aging Study (MHAS)^16^. This finding suggests that environmental factors associated with migration have a significant effect on metabolic health. Mexican immigrants to the US and their descendants are faced with significant dietary changes, including exposure to a Western diet^17^. Characterized by increased animal protein and sugar consumption with decreased complex carbohydrate consumption, Western diet contributes to pathological weight gain in diet-induced obesity^18, 19^. Importantly, host diet-derived macronutrient availability regulates the composition of the human gut microbiome^20, 21^. The gut microbiome is emerging as a powerful regulator of host physiology^22^, including metabolic function^23–25^, brain function, and behavior^26–29^. While recent studies identify key gut microbiome signatures in patients with obesity and T2D^30, 31^, the gut microbiome of individuals of Mexican descent living in the US with T2D has not been characterized in the context of its relationship to the comorbidity of AD.

The potential contribution of nongenetic factors, such as changes in the functional composition of the gut microbiome, to AD prevalence in individuals of Mexican descent with T2D is supported by several recent studies. First, a meta-analysis of two large-scale studies comparing risk factors for mild cognitive impairment (MCI) in non-Hispanic Americans *versus* Mexican Americans found that among age, education, Apolipoprotein E (APOE) ε4 status, and gender, only advanced age was a significant risk factor for MCI in Mexican Americans^32^. In a second study, analysis of serum samples identified divergent biomarker profiles in Mexican Americans with AD compared to non-Hispanic whites with AD^33^. Furthermore, Bayesian gene-set enrichment identified differential methylation in clusters of genes associated with metabolically-driven systemic inflammation in Mexican Americans with AD^34^. Additionally, there is an increase in comorbid depression in Mexican Americans with AD^4, 35^. In this context, changes in the gut microbiome, including those that alter the expression of neurotransmitters and their precursors, have been causally linked to depression in humans^36–38^. Taken together, these studies suggest that characterization of the gut microbiome signature of Americans of Mexican descent with T2D could (1) provide key mechanistic insight into the relationship between T2D and AD and (2) identify a new class of innovative treatments to prevent or delay the onset of cognitive impairment in individuals with T2D.

For this pilot study, we recruited twelve individuals of Mexican descent aged 50 – 70 years living in South Texas, within the Houston-Galveston metroplex (**Fig. 1**). Half of the study participants were subjects with T2D (sT2D) while the remaining six were healthy controls without diabetes (HC). Here we show that sT2D report a significant increase in gastrointestinal symptom severity compared to HC despite no significant difference in dietary preferences. 16S ribosomal RNA (rRNA) gene amplicon sequencing of stool specimens collected from study participants revealed only slight differences in alpha diversity and a strong interaction of BMI but not diabetes status on community structure. Moreover, we identified significant changes in the abundance of specific taxa associated with obesity and inflammation among sT2D, including a decrease in short-chain fatty acid-producing taxa *Lachnospiraceae* and *Alistipes*, as well as an increase in the known pathobiont, *Escherichia-Shigella*. Finally, predictive functional profiling identified differentially abundant gene pathways between cohorts. Together, our results suggest that alternations in the functional composition of the gut microbiome of individuals with T2D could precede and potentially contribute to risk for inflammation-associated neurodegenerative disorders.

**Figure 1.**
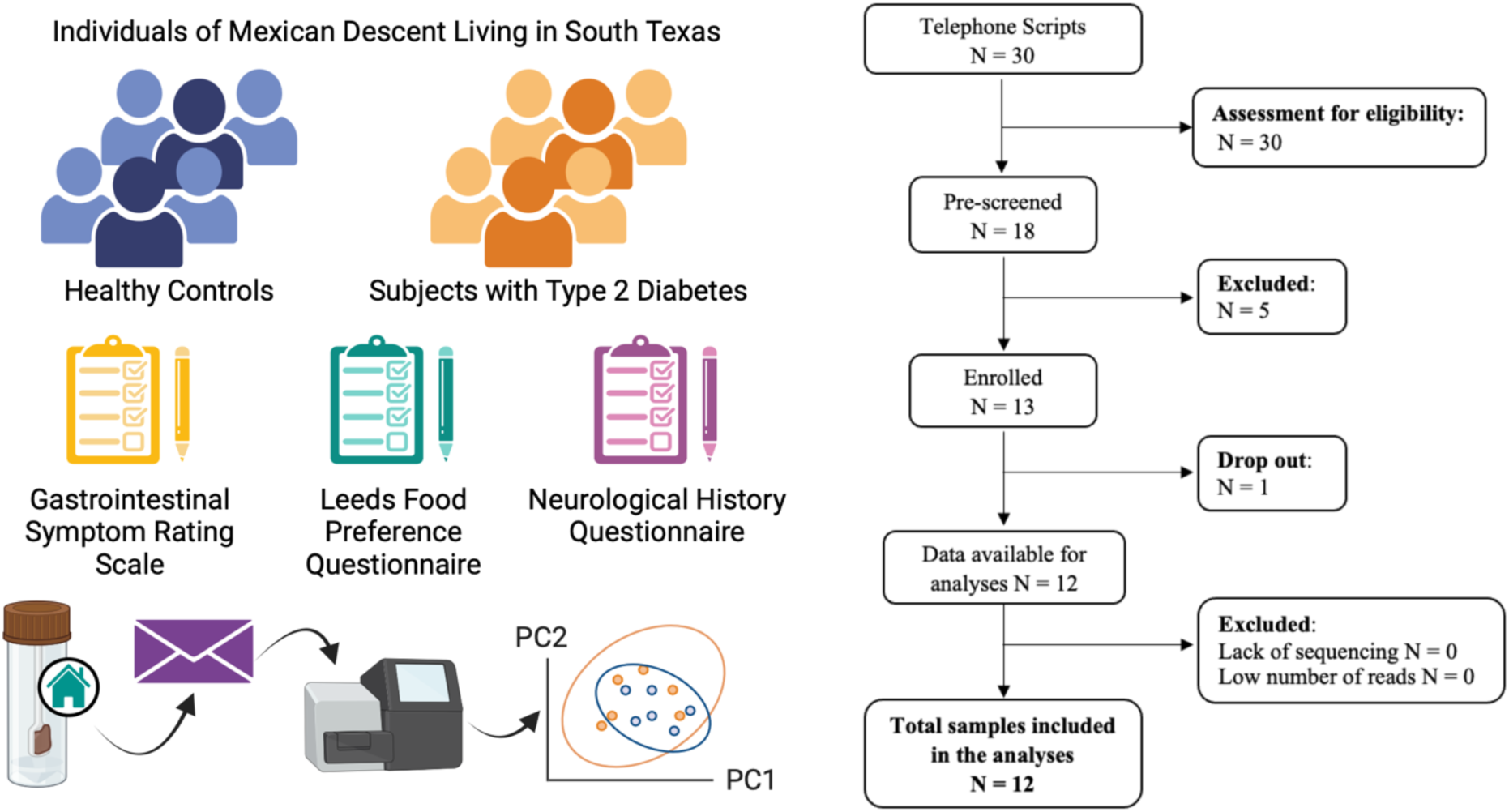
Study schematic and STORMS flowchart. Twelve individuals meeting study inclusion criteria completed the study. Each participant completed the gastrointestinal symptom rating scale (GSRS), modified Leeds Food Preference Questionnaire (LFPS), and neurological history questionnaire. Subjects submitted fecal specimens collected at home for 16S ribosomal RNA gene amplicon sequencing and analysis to characterize gut microbial communities.

## Materials and Methods

### Recruitment and Observational Clinical Study Details

We performed a case-control, cross-sectional observational clinical study based at The University of Texas Medical Branch at Galveston (UTMB). Human subjects were recruited through UTMB’s Endocrinology and Family Medicine clinics in accordance with UTMB Institutional Review Board-approved protocol 20-0201 (ClinicalTrials.gov Identifier NCT04602650; Buffington, PI; Urban, MD) between November 2020 and May 2022. Notably, the study was registered with ClinicalTrials.gov prior to participant recruitment. Subjects were pre-screened to confirm eligibility. Inclusion criteria for sT2D: 50 – 70-year-old males or females of Mexican descent living in Texas with a diagnosis of type 2 diabetes who were willing and able to give informed consent to participate in the study. Inclusion criteria for controls: 50 – 70-year-old males or females of Mexican descent living in Texas without a history of type 2 diabetes who were willing and able to give informed consent to participate in the study. Each group included English-, Spanish-speaking, and bilingual participants. Exclusion Criteria were hypertension requiring more than three anti-hypertensive medications for control, chronic kidney disease (CKD) stage 4 or higher, a history of coronary bypass or stent placement, current pregnancy, and a history of gut inflammation, including irritable bowel syndrome (IBS), celiac disease, or active diverticulitis. Pre-screened subjects that met the defined criteria were consented and enrolled at the UTMB Clinical Research Center. Participants completed medical history, dietary preference, neurological, and gastrointestinal function questionnaires (see Supplemental Information) and were instructed on home use of the provided fecal sample collection kit (DNA GenoTek OMR-200). No subjects were excluded on account of recent antibiotic usage (one subject reported taking a 10-day course of Augmentin roughly 8 weeks prior to sample collection). Subjects were compensated with two gift cards: one upon enrollment and one upon receipt of sample. Samples were de-identified and stored at –80°C until prepared for extraction and analysis. Samples were shipped on dry ice from UTMB in Galveston, TX to Baylor College of Medicine in Houston, TX where they were extracted and analyzed.

### Gastrointestinal Symptom Rating Scale (GSRS) Questionnaire and Scoring

The GSRS consists of 15 questions to assess reflux (Q2 and 3), abdominal pain (Q1, 4, and 5), indigestion (Q6–9), diarrhea (Q11, 12, and 14), and constipation (Q10, 13, and 15)^39, 40^. Subjects were asked to numerically score their subjective symptoms on a scale of 1-7 (1 = no discomfort; 7 = very severe discomfort). The sum of the scores for all 15 items is regarded as the GSRS total score. Total scores ranged from 15 (best outcome) to 105 (worst outcome). The GSRS was administered in the subject’s primary language, English or Spanish. Certified Spanish translation of each questionnaire in the study was provided by UTMB translation services. Cumulative and average scores were calculated. For the average GSRS score, averages of the five categories for subject were averaged and outliers were determined using Grubbs’ test with alpha = 0.05. Statistical significance was determined using an unpaired, two-tailed *t* test with Welch’s correction, where \**p*<0.05.

### Food Preference Questionnaire

The food preference questionnaire (FPQ)^41^ screens for known food allergies and food preferences across a variety of categories including red meat, chicken, fish, other protein (e.g., egg), grains and starches, dairy, fruit, vegetables, and sugary or fatty foods. It is a specific 3-item questionnaire in which subjects are asked to first indicate whether they identify as vegan, vegetarian, pescatarian, or none of the above. The subject is then asked whether they have any food allergies to the top eleven common food allergens and given the option to specify a food allergy not listed under, “Other.” Finally, the subject is asked to indicate preference, ranging from, “dislike a lot,” to, “like a lot,” for 59 specific food items listed in tabular format.

### Neurological History Questionnaire

The neurological questionnaire is a specific 2-item questionnaire that screens for history of neurological disorders including Alzheimer’s Disease (AD), Parkinson’s Disease (PD), Lewy Body Dementia (LBD), Bipolar Disorder, Schizophrenia, Autism Spectrum Disorder (ASD), Multiple Sclerosis (MS), Amyotrophic Lateral Sclerosis (ALS; Lou Gherig’s Disease), Guillain-Barre Syndrome (GBS), and Attention Deficit and Hyperactivity Disorder (ADHD). In the second part, subjects are asked if they have ever experienced ischemic stroke or mild traumatic brain injury (mTBI).

### Stool Sample Analysis

Metataxonomic 16S rRNA gene amplicon sequencing was performed by the Baylor College of Medicine Alkek Center for Metagenomics and Microbiome Research (CMMR) as previously described^42–44^. Briefly, bacterial genomic DNA was extracted using the DNeasy PowerSoil DNA Isolation Kit (MO BIO Laboratories, Carlsbad, CA), and the 16S ribosomal DNA (rDNA) hypervariable region 4 (V4, Forward: GTGCCAGCMGCCGCGGTAA, Reverse: GGACTACHVGGGTWTCTAAT) was amplified by PCR and sequenced on the MiSeq platform (Illumina). A bacterial mock community was used as an *in situ* positive control during extraction, amplification, and sequencing of samples. Kit elution buffers and water are used as negative controls during extraction and amplification, respectively. Raw data was uploaded into NCBI BioProject number PRJNA986954. After Trimmomatic^45^ and FastQC^46^, reads were imported into R and quality trimmed based on a minimum read length of 50, and truncated to 200– 250bp based on quality control scores < 20. Filtered reads were then ASV-inferred using DADA2^47^ followed by chimera removal and taxonomic classification using DECIPHER^48^ against the SILVA Database 138^49^. ASV counts, taxonomy, and metadata were imported into phyloseq for downstream analysis^50^. ASV counts were agglomerated to genus-level specificity, filtered based on 1% prevalence and detection, and compositionally transformed into relative abundances for taxonomic comparisons and richness estimations. Beta diversity was determined from VST-transformed ASVs based on Euclidian distance in Vegan^51^.

### Functional inference using PICRUSt2

Functional potential of the microbiome was inferred using PICRUSt2, which predicts the metagenomic content based on ASVs^52^. PICRUSt2 was used to predict functional profiles from the normalized count table, identifying functional gene families (MetaCyc and KEGG Pathways) associated with each ASV, and then summing these contributions to obtain the predicted functional profile for each sample. A comparison across diabetes status was conducted using DESeq2 with an upstream independent filtering of pathways containing less than 10 detected counts. Reported results have a log2 fold change>2.5 and an adjusted *p*-value of <0.05. Significance was corrected for multiple comparisons using the Benjamini-Hochberg multiple test correction.

## Results

### Demographics and Clinical Characteristics

Telephone scripts were read to 30 potential subjects. Of those, 18 were pre-screened. Thirteen subjects were deemed eligible and enrolled. Of those, six healthy controls without diabetes (HC; 1 male, 5 female) and six subjects with type 2 diabetes (sT2D; 3 male, 3 female) completed the study, for a total of 12 participants. The average age of HC and sT2D groups was 54.0 (HC; range 50–59) and 60.7 (sT2D; range 55–67) years. Average body mass index (BMI) was 27.9 (HC; range 21.1–34.5) and 34.6 (sT2D; range 28.5– 45.4) kg/m^2^, average A1C was 5.3 (HC; range 4.9–5.6) and 7.9 (sT2D; range 5.5–9.6) mg/dL, average systolic pressure was 117 (HC; range 97– 137) and 124 (sT2D; range 106–159) mm Hg, diastolic pressure was 72.8 (HC; range 58–87) and 79.2 (sT2D; range 72–92) mm Hg, and average estimated glomerular filtration rate (eGFR) was 88.5 (HC; range 74.5–100.0) and 113.8 (sT2D; range 78.1–165.1) mL/min/1.73 m2 (**Fig. S1**). Only age and A1C were significantly different between cohorts after FDR adjustment (**Table 1**). All sT2D were prescribed antidiabetics, including sulfonylureas (glipizide), glucagon-like peptide 1 (GLP-1) receptor agonists (semaglutide), sodium-glucose co-transporter-2 (SGLT-2) inhibitors (empagliflozin), dipeptidyl peptidase-4 (DPP-4) inhibitors (linagliptin), and biguanides (metformin) (**Table S1**). All sT2D were taking metformin. No subjects were on insulin.

**Table 1.**
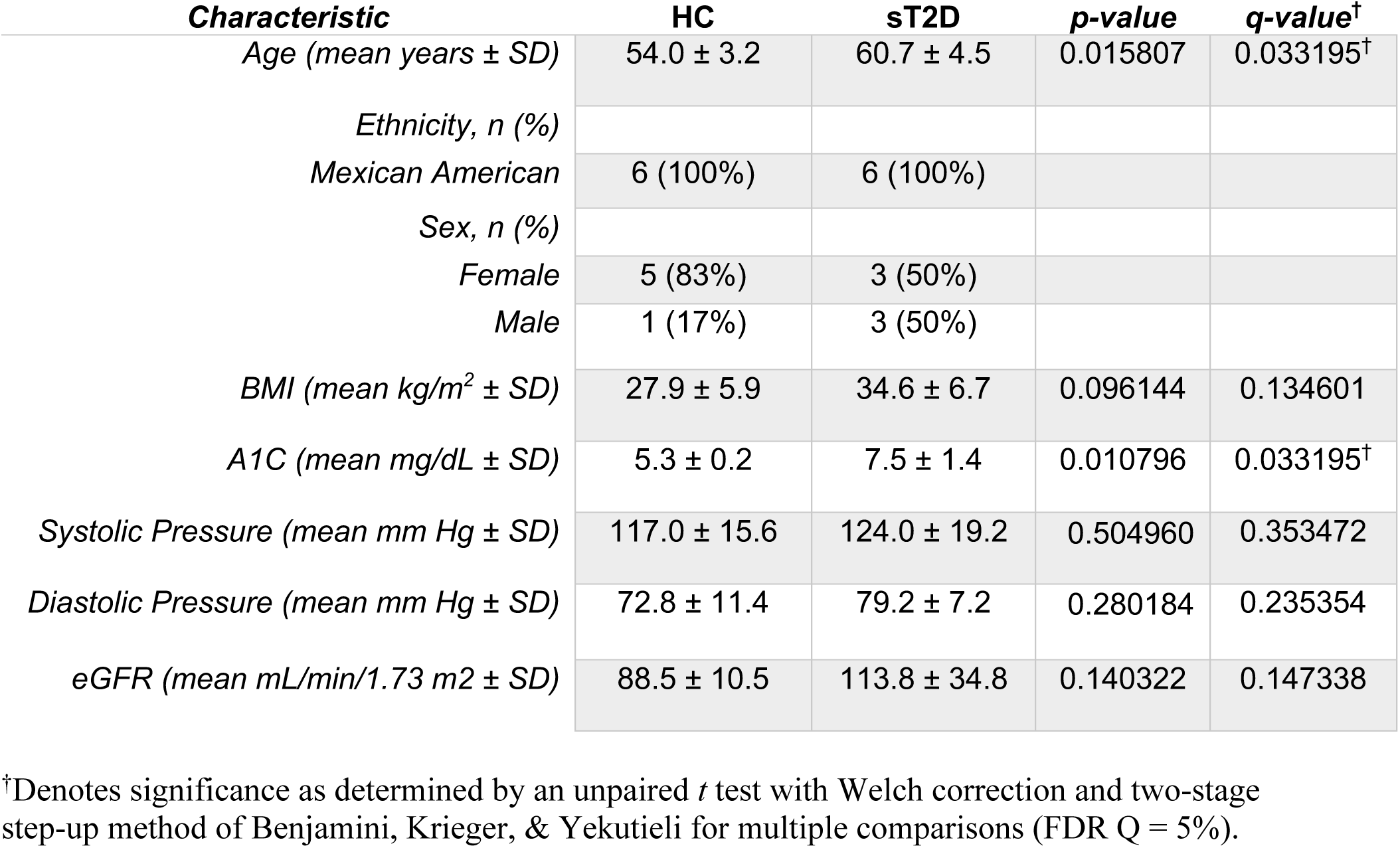
Health metrics of participants enrolled in and completing the study.

| **Characteristic** | **HC** | **sT2D** | **p-value** | **q-value<sup>†</sup>** |
| --- | --- | --- | --- | --- |
| *Age (mean years ± SD)* | 54.0 ± 3.2 | 60.7 ± 4.5 | 0.015807 | 0.033195<sup>†</sup> |
| *Ethnicity, n (%)* |  |  |  |  |
| *Mexican American* | 6 (100%) | 6 (100%) |  |  |
| *Sex, n (%)* |  |  |  |  |
| *Female* | 5 (83%) | 3 (50%) |  |  |
| *Male* | 1 (17%) | 3 (50%) |  |  |
| *BMI (mean kg/m<sup>2</sup> ± SD)* | 27.9 ± 5.9 | 34.6 ± 6.7 | 0.096144 | 0.134601 |
| *A1C (mean mg/dL ± SD)* | 5.3 ± 0.2 | 7.5 ± 1.4 | 0.010796 | 0.033195<sup>†</sup> |
| *Systolic Pressure (mean mm Hg ± SD)* | 117.0 ± 15.6 | 124.0 ± 19.2 | 0.504960 | 0.353472 |
| *Diastolic Pressure (mean mm Hg ± SD)* | 72.8 ± 11.4 | 79.2 ± 7.2 | 0.280184 | 0.235354 |
| *eGFR (mean mL/min/1.73 m<sup>2</sup> ± SD)* | 88.5 ± 10.5 | 113.8 ± 34.8 | 0.140322 | 0.147338 |

<sup>†</sup>Denotes significance as determined by an unpaired *t* test with Welch correction and two-stage step-up method of Benjamini, Krieger, & Yekutieli for multiple comparisons (FDR Q = 5%).

### Questionnaire Results: Increased gastrointestinal symptom severity, equivalent food preference reported by sT2D

No neurological events (see Materials and Methods) were reported by study participants. Interestingly, both cumulative and category-averaged GSRS scores were significantly higher in sT2D compared to HCs (**Fig. 2A, 2B**), despite no differences in dietary expressed preferences (**Fig. 2C**), suggestive of an altered gut microbial ecology in the T2D participants. No significant differences among individual GSRS categories (reflux, abdominal pain, indigestion, diarrhea, or constipation) were identified between groups (**Fig. S2**).

**Figure 2.**
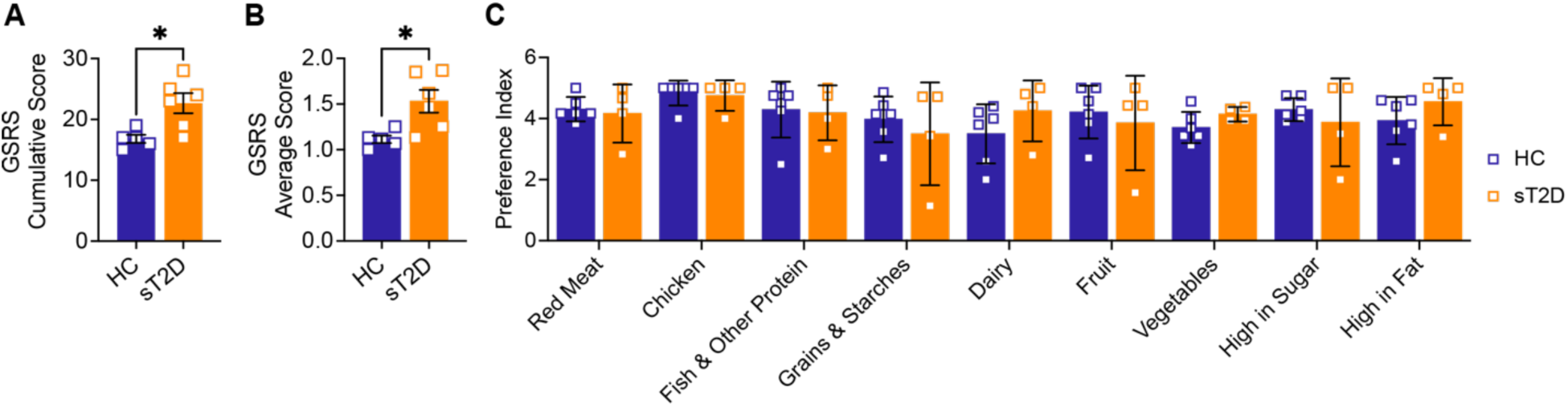
Subjects with Type 2 Diabetes report increased GI symptom severity, but have no change in dietary preferences, compared to healthy controls. (**A**) Cumulative and (**B**) average severity scores from the gastrointestinal symptom rating scale (GSRS) show that sT2D report higher GI symptom severity compared to HC [**A**: (t(6.539) = 3.305, *p* = 0.0144); **B**: (t(6.073) = 3.187, *p* = 0.0186] as determined by a two-tailed unpaired Welch’s *t*-test, with one outlier removed (Grubbs’ test; alpha = 0.05). (**C**) In contrast, food preferences are not statistically different between groups. Subjects were asked to rank foods by preference, where 0 = Not applicable, 1 = Dislike a lot, 2 = Dislike a little, 3 = Neither like nor dislike, 4 = Like a little, and 5 = Like a lot. Analysis by a mixed-effects model with Šídák’s correction for multiple comparisons did not identify any significant differences between HC and sT2D.

### Microbiome analysis reveals that BMI is a significant contributor to gut community structure

To determine if any differences were present at the genus level between sT2D and HC, we performed metataxonomic 16S ribosomal RNA (rRNA) gene amplicon sequencing of stool samples provided by study participants (see Materials & Methods). Although averages for combined alpha (within sample) diversity metrics (**Fig. 3A, C, E, G**) including amplicon sequence variants (ASVs), Chao1 index, Shannon Index, and Inverse Simpson Index were lower for sT2D compared to HC samples, they were not statistically significant. Likewise, no significant differences were observed when alpha diversity was split by sex (**Fig. 3B, D, F, H**). Although GSRS data suggested a disturbance to gut community structure of sT2D, no significant differences in beta (between sample) diversity were observed as measured by permutational ANOVA of Bray-Curtis (**Fig. 3I**), and Weighted (Diabetes Status: p = 0.5682, R^2^ = 0.0904, permutations = 9999; BMI: p = 0.8771, R^2^ = 0.0159, permutations = 9999) or Unweighted UniFrac distances (Diabetes Status: p = 0.6119, R^2^ = 0.0780, permutations = 9999; BMI: p = 0.0121, R^2^ = 0.1470, permutations = 9999), possibly due to the small sample size of this pilot cohort. However, sample clusters did trend towards significance based on body mass index (BMI) values (**Fig. 3J**). Specifically, use of the Envfit function to fit environmental vectors age and BMI onto an ordination plot revealed that BMI, but not age, is a significant driver of community structure (**Fig. S3**).

**Figure 3.**
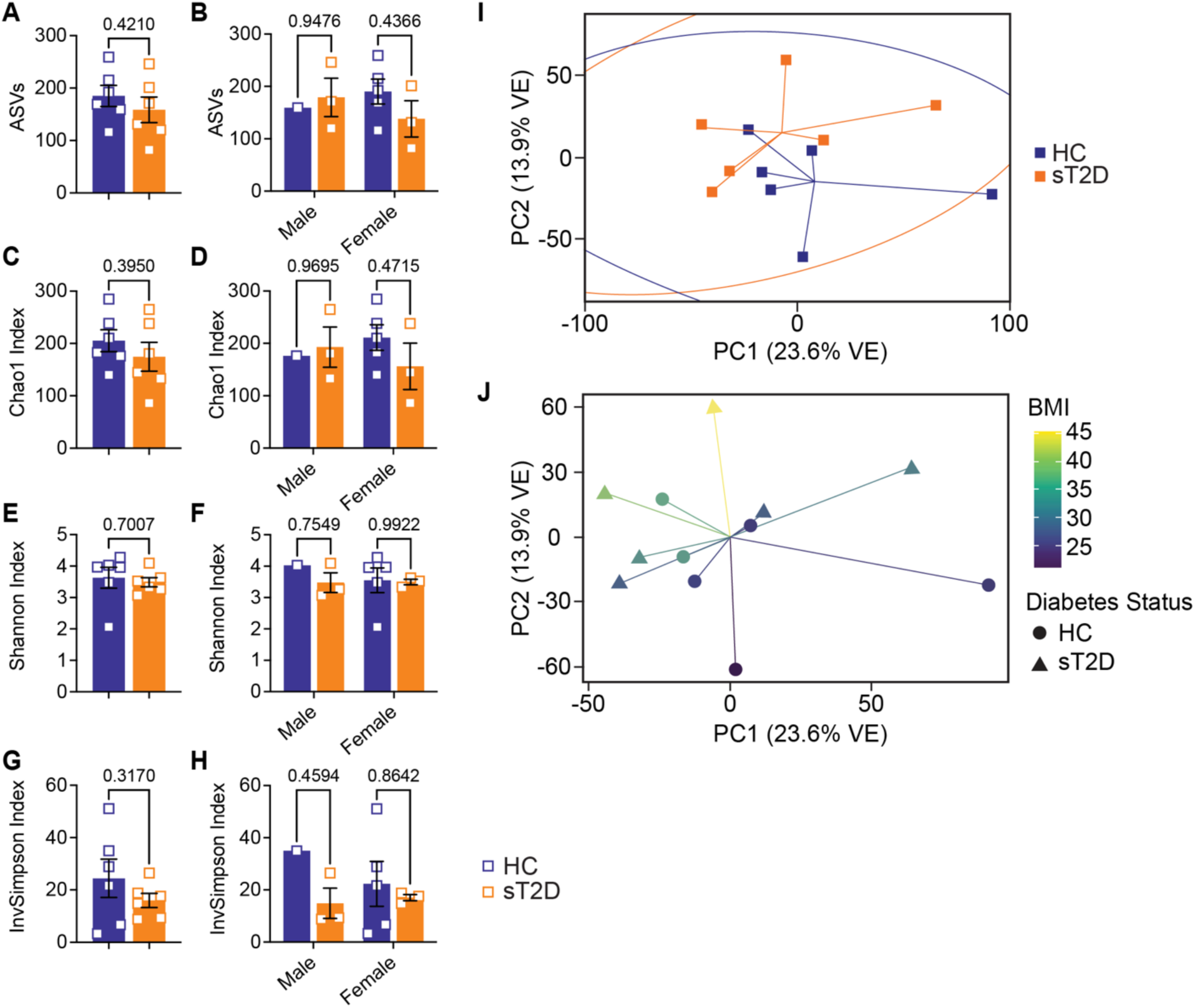
Analysis of alpha diversity metrics among study participants revealed a trending decrease in microbial diversity among subjects with T2D. (**A, C, E, G**) Alpha diversity graphs for males and females combined analyzed by two-tailed, unpaired Welch’s t-test: (**A**) ASVs (*t*(9.661) = 0.8404, p = 0.4210), (**C**) Chao1 Index (*t*(9.323) = 0.8917, p = 0.395), (**E**) Shannon Index (*t*(6.877) = 0.4008, p = 0.7007), and (**G**) Inverse Simpson Index (*t*(6.328) = 1.086, p = 0.317). (**B, D, F, H**) Alpha diversity graphs separated by sex and analyzed by two-way ANOVA with Šídák’s correction for multiple comparisons: (**B**) ASVs (Male: *t*(8) = 0.3010, p = 0.9476; Female: *t*(8) = 1.242, p = 0.4366), (**D**) Chao1 Index (Male: *t*(8) = 0.2280, p = 0.9695; Female: *t*(8) = 1.177, p = 0.4715), (**F**) Shannon Index (Male: *t*(8) = 0.6981, p = 0.7549; Female: *t*(8) = 0.1142, p = 0.9922), and (**H**) Inverse Simpson Index (Male: *t*(8) = 1.199, p = 0.4594; Female: *t*(8) = 0.4986, p = 0.8642). Principal coordinate analysis of (**I**) Bray-Curtis dissimilarities analyzed by permutational ANOVA (p = 0.175, R^2^ = 0.1028, permutations = 999) did not reveal statistically significant clusters between subject groups but did trend toward significance based on (**J**) BMI values (p = 0.0740, R^2^ = 0.1188, permutations = 999). Similarly, unweighted UniFrac (which incorporates phylogenetic distance but does not consider abundance) analysis was significantly different for BMI but not Diabetes Status; however, this significance was not observed by Weighted UniFrac (which does consider abundance). Importantly, betadisper analysis did not reveal significant *within group* significant differences by either method (Bray-Curtis: p = 0.7844; Unweighted and Weighted UniFrac: p = 0.1512 and 0.9136, respectively).

### Microbiome analysis reveals significant differences in abundance of seventeen taxa

We next used linear discriminant analysis (LDA) effect size (LEfSe)^53^ to determine which taxa were most likely to explain differences between HC and sT2D cohorts and identified seventeen differentially abundant taxa (**Fig. 4A**). sT2D samples were enriched for *Clostridum innocuum*, *Streptococcus*, *Enterobacter*, and *Escherichia-Shigella* (**Fig. 4B–E**), while HC samples were enriched with genera from *Lachnospiraceae*, *Faecalibacterium*, and *Alistipes*, among others (**Fig. 4F–R**).

**Figure 4.**
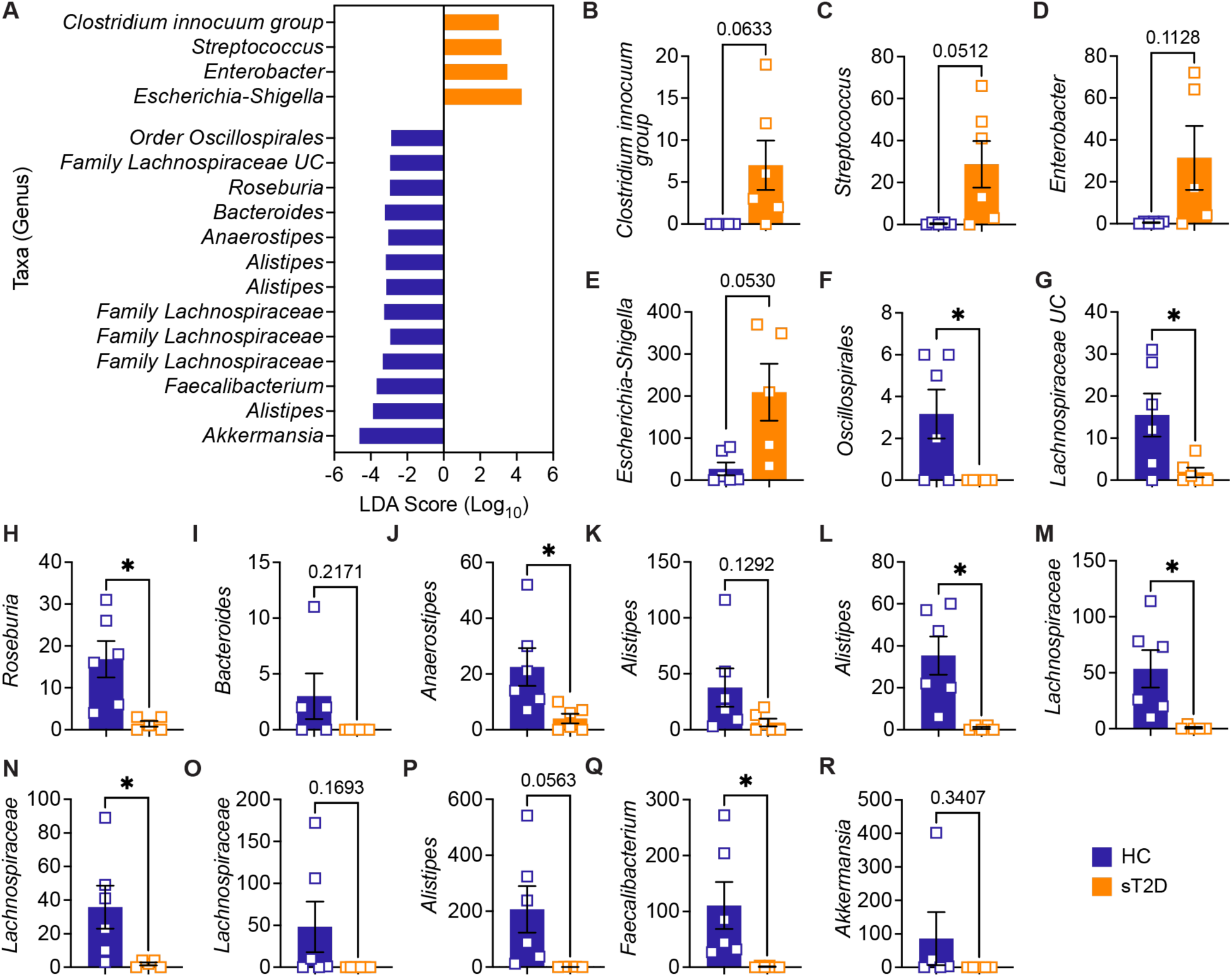
Linear Discriminant Analysis (LDA) Effect Size (LEfSe) detected significant abundance differences in taxa between healthy controls and subjects with Type 2 Diabetes. (**A**) Histogram of LDA scores (Log_10_) for genus-level taxa with p < 0.05. UC; uncultured. Group comparisons of ASVs for (**B**) *Clostridium innocuum group* (*t*(5) = 2.378, p = 0.0633), (**C**) *Streptococcus* (*t*(5.005) = 2.550, p = 0.0512), (**D**) *Enterobacter* (*t*(4.002) = 2.025, p = 0.1128), and (**E**) *Escherichia-Shigella* (*t*(4.403) = 2.627, p = 0.2062), (**F**) *Oscillospirales* (*t*(5) = 2.714, p = 0.0421), (**G**) *Lachnospiraceae UC* (*t*(5.492) = 2.604, p = 0.0439), (**H**) *Roseburia* (*t*(5.242) = 3.502, p = 0.016), (**I**) *Bacteroides* (*t*(4) = 1.464, p = 0.2171), (**J**) *Anaerostipes* (*t*(5.652) = 2.649, p = 0.0403), (**K**) *Alistipes* (*t*(5.399) = 1.789, p = 0.1292), (**L**) *Alistipes* (*t*(5.029) = 3.787, p = 0.0127), (**M**) *Lachnospiraceae* (*t*(5.023) = 3.144, p = 0.0254), (**N**) *Lachnospiraceae* (*t*(5.049) = 2.631, p = 0.046), (**O**) *Lachnospiraceae* (*t*(5) = 1.606, p = 0.1693), (**P**) *Faecalibacterium* (*t*(5.002) = 2.608, p = 0.0478), (**Q**) *Alistipes* (*t*(5) = 2.474, p = 0.0563), (**R**) *Akkermansia* (*t*(4) = 1.081, p = 0.3407), as determined by an unpaired, two-tailed Welch’s *t*-test.

### Predictive functional profiling identifies differentially abundant gene pathways between cohorts

Finally, to perform predictive functional profiling, we first used PICRUSt2 to generate functional outputs based on Kyoto Encyclopedia of Genes and Genomes (KEGG)^54, 55^ and MetaCyc databases (**Tables 2 and 3**, **Fig. 5**), and then performed pairwise comparisons using DESeq2 to determine if any of these pathways were significantly enriched in sT2D compared to HC samples. We identified four significantly differentially abundant KEGG pathways (**Table 2**): Bacterial invasion of epithelial cells, *Staphylococcus aureus* infection, alpha-linolenic acid metabolism, and polycyclic aromatic hydrocarbon degradation, and 15 significantly differentially abundant MetaCyc metabolic pathways (**Table 3**, **Figure 5**).

**Figure 5.**
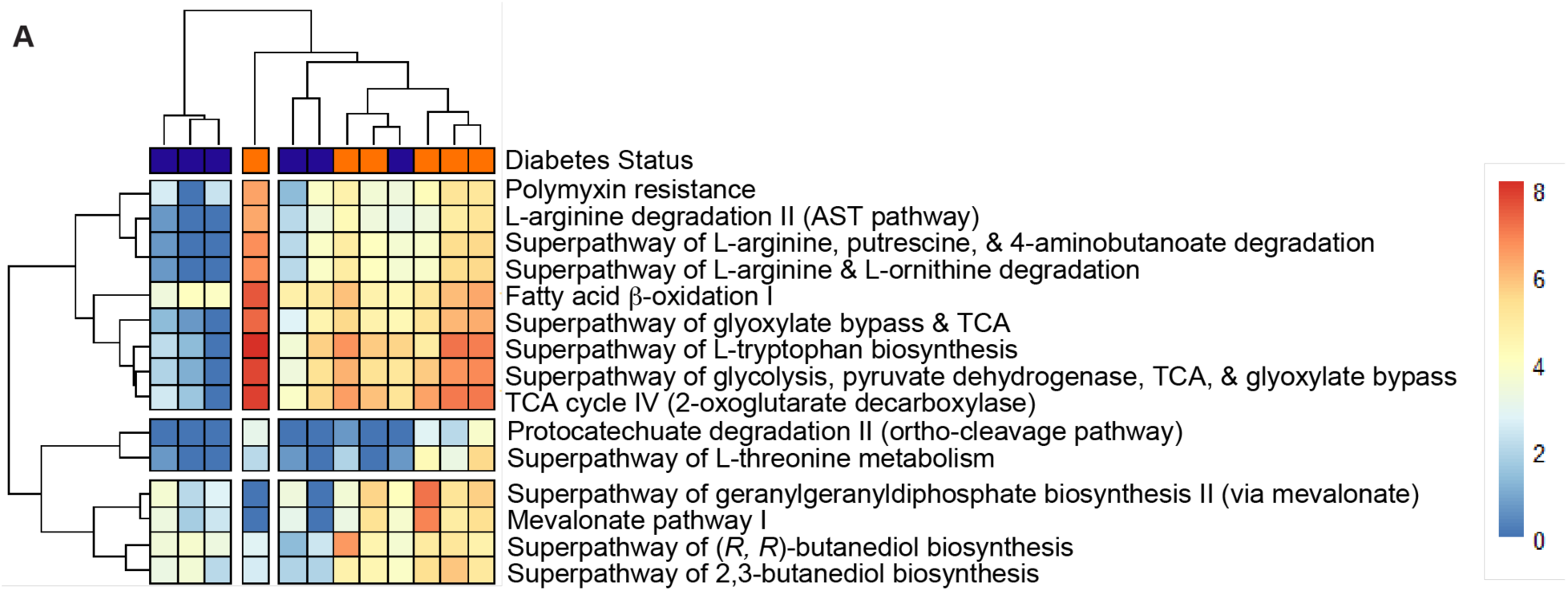
Heatmap of statistically significant Metacyc features that differ between groups with p < 0.05. (**A**) Heatmap based on PICRUSt2 output (See Materials and Methods and Table 2 for p-values).

**Table 2.** Results of four significant differentially abundant ASV-inferred KEGG pathways associated with sT2D compared to HC samples.

| **Feature** | **Pathway Name** | **Adjusted p-value** |
| --- | --- | --- |
| **KO00592** | Alpha-linolenic acid metabolism | 0.0066 |
| **KO00624** | Polycyclic aromatic hydrocarbon degradation | 0.0034 |
| **KO05150** | Staphylococcus aureus infection | 0.0001 |
| **KO05100** | Bacterial invasion of epithelial cells | 0.0074 |

**Table 3.** Results of 15 significant differentially abundant ASV-inferred MetaCyc metabolic pathways associated with sT2D compared to HC samples.

| **Pathway** | **Description** | **Log<sub>2</sub>FoldChange** | **Adj. p-value** |
| --- | --- | --- | --- |
| **ARGDEG-PWY** | Superpathway of L-arginine, putrescine, & 4-aminobutanoate degradation | 3.887 | 0.049 |
| **AST-PWY** | L-arginine degradation II | 3.919 | 0.049 |
| **FAO-PWY** | Fatty acid β-oxidation I | 2.806 | 0.019 |
| **GLYCOLYSIS-TCA-GLYOX-BYPASS** | Superpathway of glycolysis, pyruvate dehydrogenase, TCA, & glyoxylate bypass | 3.561 | 0.036 |
| **ORNARGDEG-PWY** | Superpathway of L-arginine & L-ornithine degradation | 3.887 | 0.049 |
| **P105-PWY** | TCA cycle IV (2-oxoglutarate decarboxylase) | 3.686 | 0.029 |
| **P125-PWY** | Superpathway of (*R,R*)-butanediol biosynthesis | 2.873 | 0.048 |
| **PROTOCATECHUATE-ORTHOCLEAVAGE-PWY** | Protocatechuate degradation II (ortho-cleavage pathway) | 6.595 | 0.049 |
| **PWY-5910** | Superpathway of geranylgeranyldiphosphate biosynthesis I (via mevalonate) | 3.911 | 0.049 |
| **PWY-6396** | Superpathway of 2,3-butanediol biosynthesis | 2.865 | 0.049 |
| **PWY-6629** | Superpathway of L-tryptophan biosynthesis | 3.319 | 0.049 |
| **PWY-922** | Mevalonate pathway I | 3.978 | 0.049 |
| **PWY0-1338** | Polymyxin resistance | 3.374 | 0.049 |
| **TCA-GLYOX-BYPASS** | Superpathway of glyoxylate bypass & TCA | 3.750 | 0.036 |
| **THREOCAT-PWY** | Superpathway of L-threonine metabolism | 7.310 | 0.014 |

## Discussion

Our study adds to a growing body of literature linking inflammation and dysbiosis of the gut microbiome to AD susceptibility (for review see ^56^). For instance, a recent study of a small cohort of dementia patients revealed divergent gut microbiota composition, increased gut permeability, and inflammation, implicating a microbial determinant in neuroimmune dysregulation, an emerging player in neurodegeneration^57^. The aim of this study was to examine if changes in gut community structure occur in individuals with T2D compared to individuals without diabetes in a population at increased risk for AD. This multidisciplinary pilot study builds on previous investigations into the gut microbial community structure of a similar cohort of Americans of Mexican descent with high rates of obesity and diabetes^11, 12^. Although not statistically significant, our finding here that some metrics of within-sample (alpha) diversity were slightly lower in gut microbiomes of sT2D compared to HC (**Fig. 3A–H**) agrees with a previous study reporting altered composition and functional capacity of gut microbiomes in obese patients^58^. This same study also reported differences in between-sample (beta) diversity, suggesting that a larger sample size may detect differences not captured in our small pilot study. Importantly, however, our findings agree with other previously reported associations between T2D and changes in abundance of specific taxa, including over-representation of inflammation-promoting taxa (*Escherichia-Shigella*, *Enterobacter*, and *Clostridium-innocuulum*), and underrepresentation of short-chain fatty acid (specifically, butyrate)-producing taxa (*Alistipes*, *Faecalibacterium*, and *Lachnospiraceae*) (**Fig. 4**)^58^. Lastly, although not statistically significant, the greater disparity in alpha diversity metrics that we observed between sT2D and HC females (reflected by ASVs and Chao1 index only) may still be physiologically relevant, given that 1) women are disproportionately affected by AD and 2) recently published results in preclinical animal model experiments performed by our lab showing a sexually dimorphic impact of diet and supplementation (probiotics) alike on host gut microbiota composition^44^. Given the person-to-person variability observed among gut microbiome profiles of aging populations^59^, our results from a small cohort could be indicative of a predictive disease signature among patients with T2D predisposed toward AD.

Our study is not without limitations. First, enrollment took place from 2020–2022, and recruitment challenges associated with the COVID-19 pandemic resulted in a small sample size (N =12). Longitudinal, sufficiently-powered studies are needed to continue exploring how alterations in gut microbiome community structure contribute to neurodegeneration. Second, while we did not detect any differences in food preferences between HC and sT2D, it is possible that the food preference questionnaire was not ideally suited to our distinct cohort (Americans of Mexican descent living in South Texas). Additionally, all six sT2D were prescribed metformin, a known modulator of gut microbiome community composition^60^, and treatment with sulfonylurea drugs (taken by one subject here) increases risk for hypoglycemia, which may accelerate dementia^61^. Notably, Thingholm *et al*. found clear associations between host microbiome variation and medications, supplementations, and diet, making direct changes due to diabetes status difficult to establish. Therefore, the potentially confounding variable of medication should be considered in subsequent studies examining links between the gut microbiome, diabetes, and dementia. Also of note, in our study BMI was not significantly different between cohorts (*p* = 0.096144, **Table 1**) but did strongly associated with community structure (**Table 1**, **Fig. 3, Fig. S3**). Notably, it is difficult to match BMI between cohorts in a diabetes study, given that a BMI ≥ 30 kg/m^2^ has been shown to be a strong indicator of “adult lifetime risk of diabetes^62^;” nonetheless, the role of diabetes specifically (independent of obesity) as a driver of AD must be carefully teased out in future work. Finally, metataxonomic sequencing is of limited (genus/ASV) resolution, whereas metagenomic whole genome shotgun sequencing of participant samples would provide species-to-strain level-specific information as well as functional data (as opposed to predicted functions which may not capture “rare environment-specific functions^52^” of bacteria whose genomes are not currently well-represented). Indeed, metagenomic combined with metatranscriptomic analysis of gut microbiota from patients with T2D both pre– and post-medication, and post-AD diagnosis, will be critical to differentiate between changes in the composition and functional profile of the gut microbiota due to various medications and those that may directly contribute to neurodegeneration.

Data presented here highlight the potential of such studies to provide mechanistic insight into how T2D predisposes individuals to the neuropathology and cognitive impairment characteristic of AD. Given our findings that sT2D reported increase gastrointestinal symptoms compared to HC (**Fig. 2**), a noteworthy ASV-inferred functional KEGG ortholog (KO) differentially expressed in sT2D in our study is the bacterial invasion of epithelial cells (KO05100, **Table 2**). Additionally, given Thus, future studies examining gut permeability and gut barrier integrity in T2D patient populations predisposed to AD may be warranted. Conversely, the microbiome is also a source of neuroprotective metabolites, such as indole derivatives which modulate host inflammation^63^. Surprisingly, differentially abundant ASV-inferred MetaCyc metabolic pathways (including the tryptophan pathway, an indole precursor) were all elevated in sT2D compared to HC (**Table 3**, **Fig. 5**); however, this insight might identify a novel class of interventions that target the host gut microbiome for the prevention of early onset AD in a predisposed population, as well as the population at large. This study brings us one step closer to the identification of a microbial or microbially-associated signature that predicts dementia risk, as well as pre– or probiotics that can modulate said signatures, which would be a key breakthrough that could revolutionize care for patients with metabolic and neurodegenerative disorders alike. Moreover, it highlights the therapeutic potential of targeting the gut microbiome to dampen neuroinflammation in the context of metabolic and neurodegenerative disorders.

## Data Availability

Deidentified primary data will be made available upon request. The BioProject associated with this study where sequence data can be found is PRJNA986954. R code is available at https://github.com/MADscientist314/lmatz. A STORMS (Strengthening The Organizing and Reporting of Microbiome Studies)^64^ checklist is available at 10.5281/zenodo.10723337.

## Acknowledgements

We would like to thank Kathleen Randolph, Christopher Danesi, Lori Simon, Lisa Hernandez Garcia, and Heidi Sperger for their invaluable clinical/IRB knowledge and guidance during this project, and Ruby Izar-Shea and Debbie Love for their medical translation services. We would also like to thank the UTMB Endocrinology and Family Medicine clinicians including Drs. Ruba Riachy, Cindy Lyou Chan, Randall Urban, Nisarg Shah, Syed Azhar, Oyetokunbo Ibidapo-Obe, Laura Porterfield, Michael Allen, and Zuleica Santiago-Delgado who assisted in recruiting patients for this study, as well as the patients who participated. This study was conducted with the support of the Institute for Translational Sciences at UTMB, supported in part by a Clinical & Translational Science Award (UL1TR001439) from the National Center for Research Resources, now at the National Center for Advancing Translational Sciences, NIH. Salary support for LM during a portion of the project was provided by NIH 1T32 AG067952-01 (UTMB Mitchell Center for Neurodegenerative Diseases). We would like to thank the Alkek Center for Metagenomics & Microbiome Research at BCM for RNA extraction, 16S rRNA gene amplicon sequencing, and data analysis platform Atima2 (https://atima.research.bcm.edu). Data were visualized and analyzed using GraphPad Prism version 8.4.3 for Mac OS X, GraphPad Software, San Diego, California USA, www.graphpad.com. Figures were designed in Adobe Illustrator and BioRender.

## Supplemental Information

**Figure S1.**
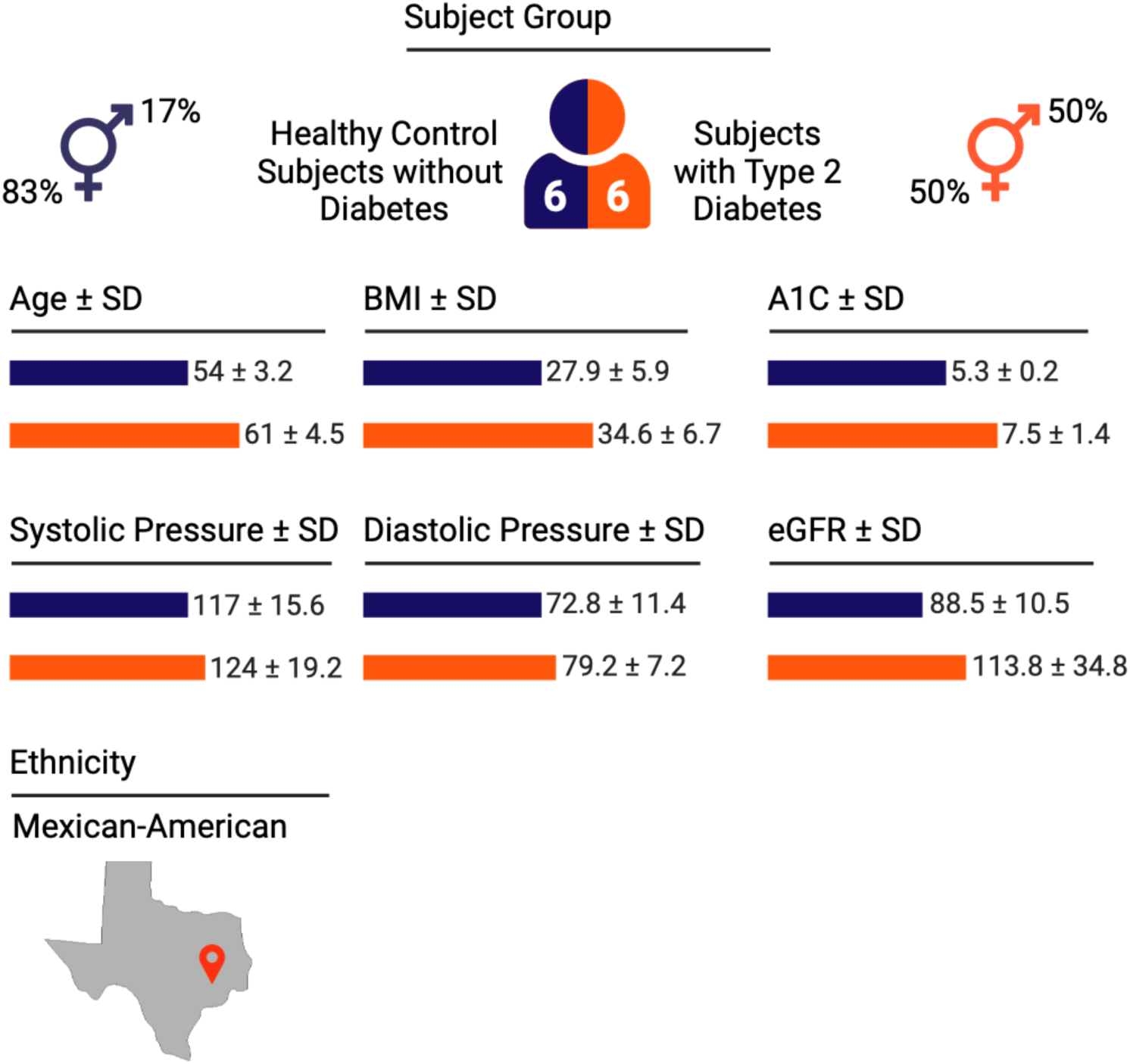
Study subject demographic schematic. Demographics of healthy controls without diabetes and subjects with type 2 diabetes as reported in Table 1. SD, standard deviation of the mean. Related to **Figure 1**, **Table 1**.

**Figure S2.**
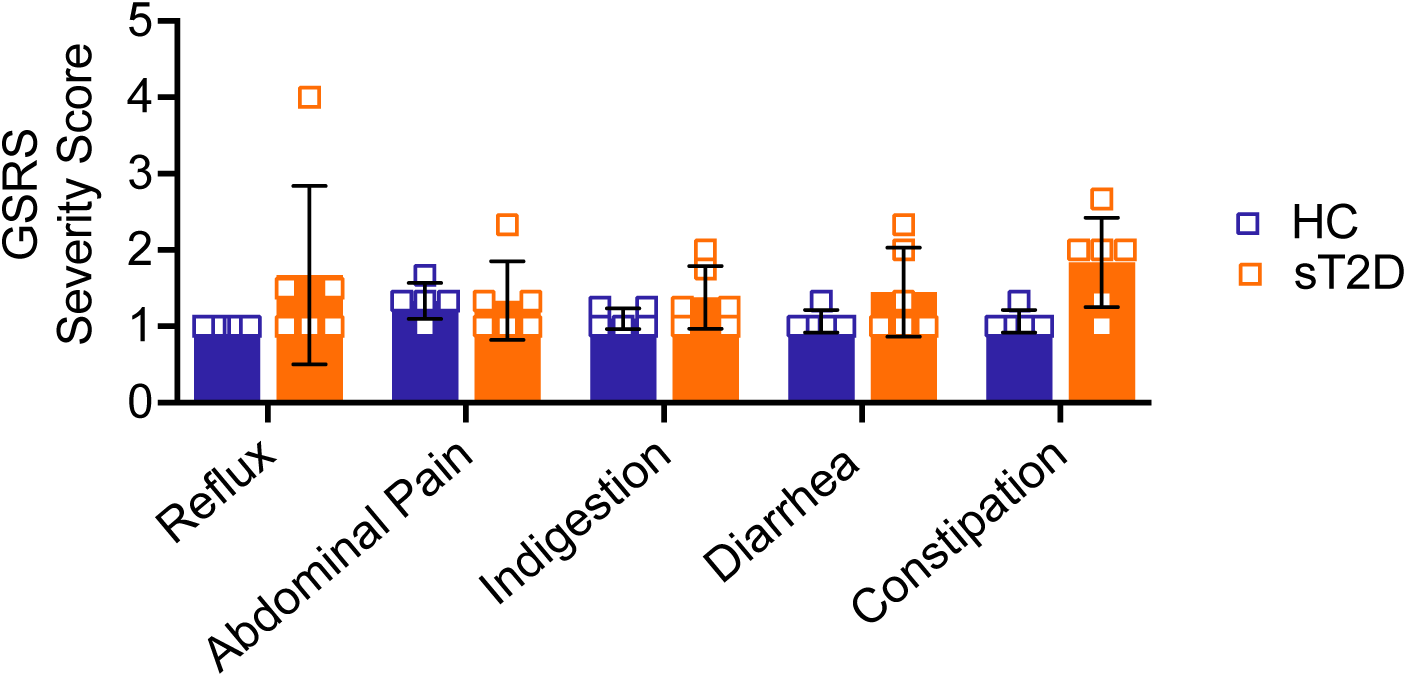
Gastrointestinal Symptom Rating Scale scores averaged by category. Average severity scores (y-axis) for each gastrointestinal symptom category (X-axis) did not differ significantly between HC and sT2D as determined by two-way ANOVA [Reflux: (t(45) = 2.055, *p* = 0.2084); Abdominal Pain: (t(45) = 1.028e-9, *p* = >0.9999.); Indigestion: (t(45) = 0.8478, *p* = 0.9229); Diarrhea (t(45) = 1.165, *p* = 0.7631); Constipation (t(45) = 2.364, *p* = 0.1074)]. Related to **Figure 2**.

**Figure S3.**
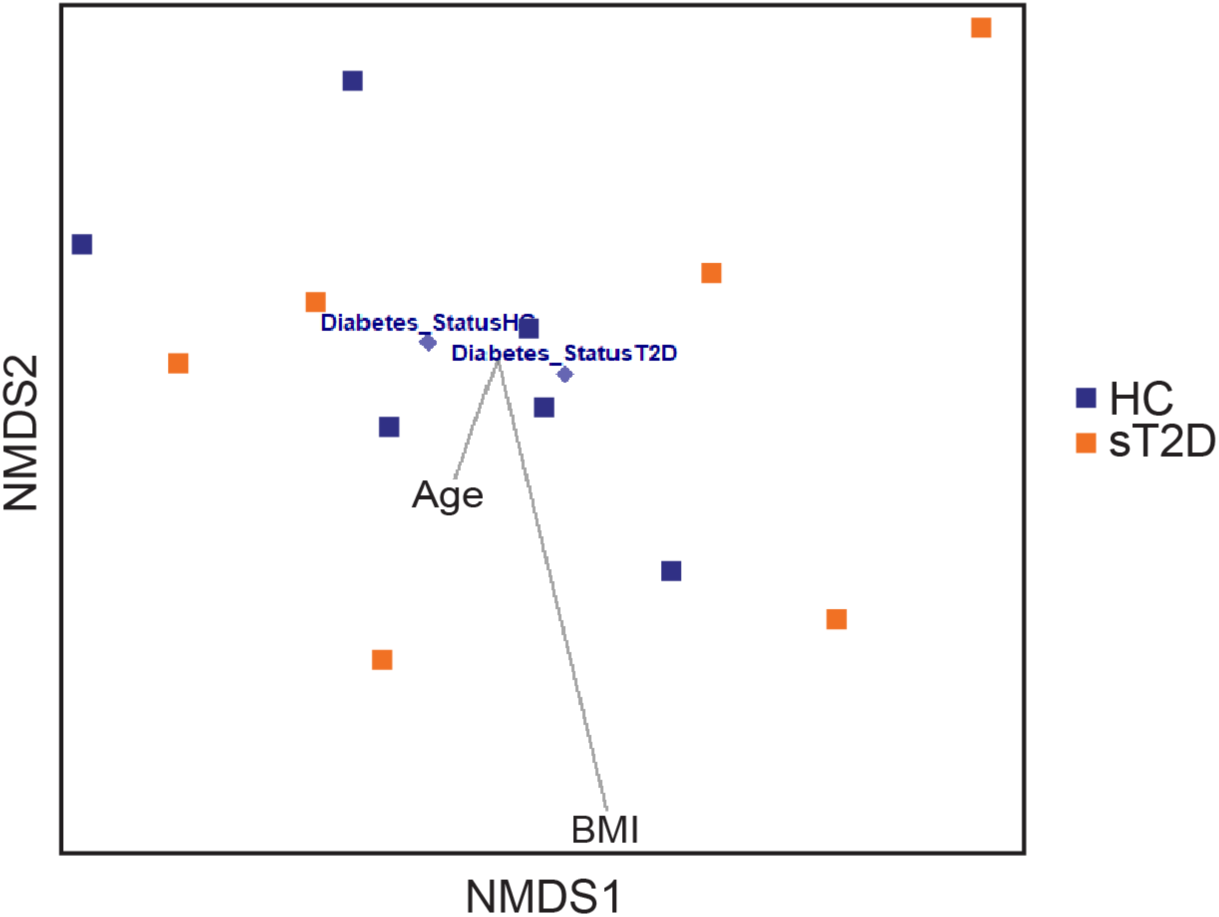
Non-metric multidimensional scaling (NMDS) plot with Envfit overlay comparing HC and sT2D samples revealed that BMI, but not age, is a strong variable driving community structure. Envfit plot incorporating vectors of BMI and Age (BMI: p = 0.007, R^2^ = 0.7062, permutations = 999; Age: p = 0.750, R^2^ = 0.0548, permutations = 999). Bray–Curtis dissimilarity was used as the distance metric for the overlay of BMI and age vectors. HC and sT2D samples shown in blue and orange, respectively. Related to **Figures 3** and **S1**.

**Table S1.** Categories of select drugs prescribed to subjects with Type 2 Diabetes.

| **Drug Category** | **# sT2D using** |
| --- | --- |
| Anti-hypertensives | 4/6 |
| Anti-diabetics | 6/6 |
| Anti-inflammatories | 3/6 |
| PPIs/H2As/Anti-histamines | 4/6 |
| Anti-depressants, anti-convulsants | 2/6 |
| Anti-lipemics | 4/6 |

## Questionnaires

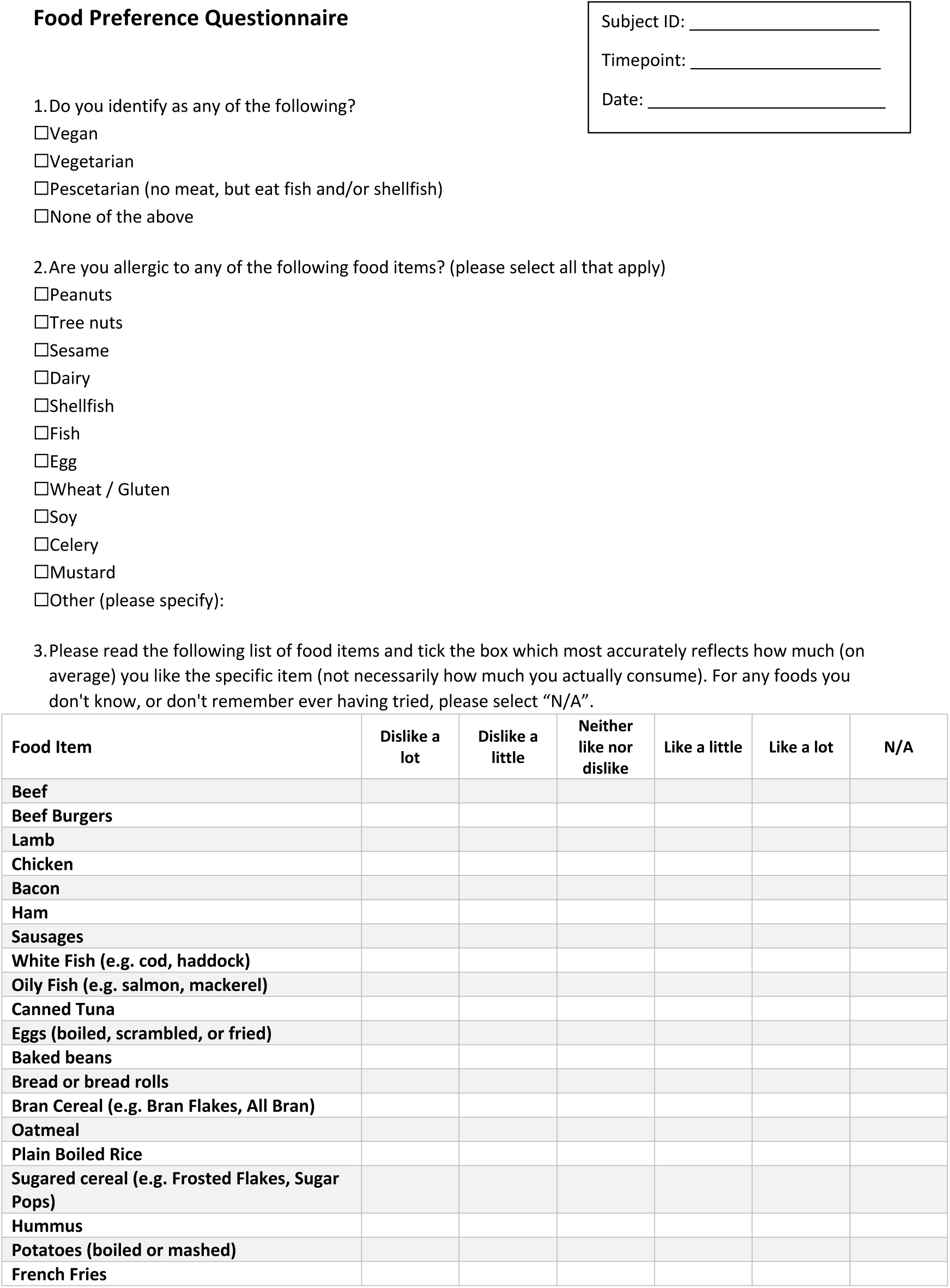
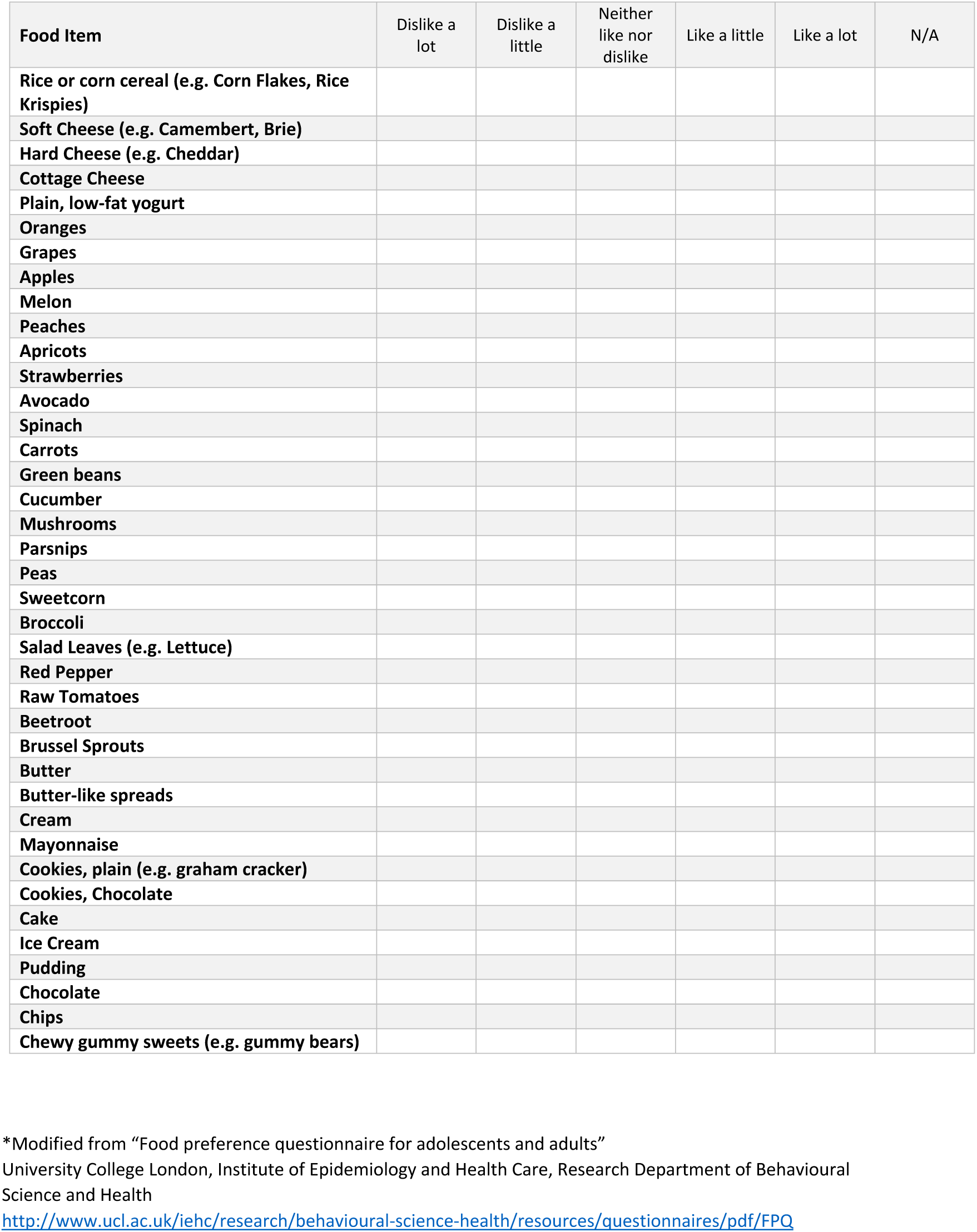
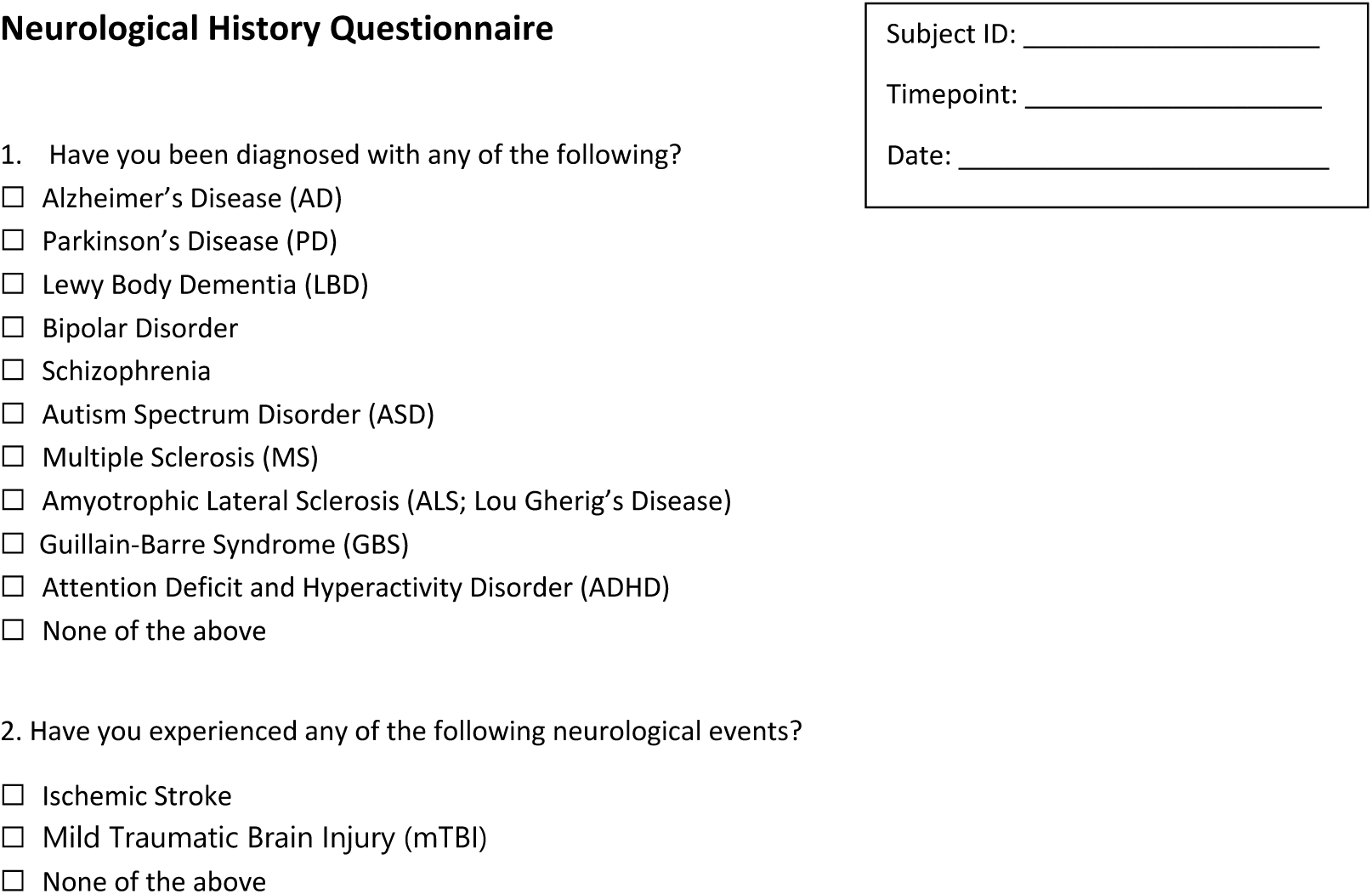

## THE GASTROINTESTINAL SYMPTOM RATING SCALE (GSRS)

Please read this first:

This survey contains questions about how you have been feeling and what it has been like DURING THE PAST WEEK. Mark the choice that best applies to you and your situation with an “X” in the box.

1. Have you been bothered by PAIN OR DISCOMFORT IN YOUR UPPER ABDOMEN OR THE PIT OF YOUR STOMACH during the past week?

⎕ No discomfort at all
⎕ Minor discomfort
⎕ Mild discomfort
⎕ Moderate discomfort
⎕ Moderately severe discomfort
⎕ Severe discomfort
⎕ Very severe discomfort
2. Have you been bothered by HEARTBURN during the past week? (By heartburn we mean an unpleasant stinging or burning sensation in the chest.)

⎕ No discomfort at all
⎕ Minor discomfort
⎕ Mild discomfort
⎕ Moderate discomfort
⎕ Moderately severe discomfort
⎕ Severe discomfort
⎕ Very severe discomfort
3. Have you been bothered by ACID REFLUX during the past week? (By acid reflux we mean the sensation of regurgitating small quantities of acid or flow of sour or bitter fluid from the stomach up to the throat.)

⎕ No discomfort at all
⎕ Minor discomfort
⎕ Mild discomfort
⎕ Moderate discomfort
⎕ Moderately severe discomfort
⎕ Severe discomfort
⎕ Very severe discomfort
4. Have you been bothered by HUNGER PAINS in the stomach during the past week? (This hollow feeling in the stomach is associated with the need to eat between meals.)

⎕ No discomfort at all
⎕ Minor discomfort
⎕ Mild discomfort
⎕ Moderate discomfort
⎕ Moderately severe discomfort
⎕ Severe discomfort
⎕ Very severe discomfort
5. Have you been bothered by NAUSEA during the past week? (By nausea we mean a feeling of wanting to throw up or vomit.)

⎕ No discomfort at all
⎕ Minor discomfort
⎕ Mild discomfort
⎕ Moderate discomfort
⎕ Moderately severe discomfort
⎕ Severe discomfort
⎕ Very severe discomfort
6. Have you been bothered by RUMBLING in your stomach during the past week? (Rumbling refers to vibrations or noise in the stomach.)

⎕ No discomfort at all
⎕ Minor discomfort
⎕ Mild discomfort
⎕ Moderate discomfort
⎕ Moderately severe discomfort
⎕ Severe discomfort
⎕ Very severe discomfort
7. Has your stomach felt BLOATED during the past week? (Feeling bloated refers to swelling often associated with a sensation of gas or air in the stomach.)

⎕ No discomfort at all
⎕ Minor discomfort
⎕ Mild discomfort
⎕ Moderate discomfort
⎕ Moderately severe discomfort
⎕ Severe discomfort
⎕ Very severe discomfort
8. Have you been bothered by BURPING during the past week? (Burping refers to bringing up air or gas from the stomach via the mouth, often associated with easing a bloated feeling.)

⎕ No discomfort at all
⎕ Minor discomfort
⎕ Mild discomfort
⎕ Moderate discomfort
⎕ Moderately severe discomfort
⎕ Severe discomfort
⎕ Very severe discomfort
9. Have you been bothered by PASSING GAS OR FLATUS during the past week? (Passing gas or flatus refers to the need to release air or gas from the bowel, often associated with easing a bloated feeling.)

⎕ No discomfort at all
⎕ Minor discomfort
⎕ Mild discomfort
⎕ Moderate discomfort
⎕ Moderately severe discomfort
⎕ Severe discomfort
⎕ Very severe discomfort
10. Have you been bothered by CONSTIPATION during the past week? (Constipation refers to a reduced ability to empty the bowels.)

⎕ No discomfort at all
⎕ Minor discomfort
⎕ Mild discomfort
⎕ Moderate discomfort
⎕ Moderately severe discomfort
⎕ Severe discomfort
⎕ Very severe discomfort
11. Have you been bothered by DIARRHEA during the past week? (Diarrhea refers to a too frequent emptying of the bowels.)

⎕ No discomfort at all
⎕ Minor discomfort
⎕ Mild discomfort
⎕ Moderate discomfort
⎕ Moderately severe discomfort
⎕ Severe discomfort
⎕ Very severe discomfort
12. Have you been bothered by LOOSE STOOLS during the past week? (If your stools (motions) have been alternately hard and loose, this question only refers to the extent you have been bothered by the stools being loose.)

⎕ No discomfort at all
⎕ Minor discomfort
⎕ Mild discomfort
⎕ Moderate discomfort
⎕ Moderately severe discomfort
⎕ Severe discomfort
⎕ Very severe discomfort
13. Have you been bothered by HARD STOOLS during the past week? (If your stools (motions) have been alternately hard and loose, this question only refers to the extent you have been bothered by the stools being hard.)

⎕ No discomfort at all
⎕ Minor discomfort
⎕ Mild discomfort
⎕ Moderate discomfort
⎕ Moderately severe discomfort
⎕ Severe discomfort
⎕ Very severe discomfort
14. Have you been bothered by an URGENT NEED TO HAVE A BOWEL MOVEMENT during the past week? (This urgent need to go to the toilet is often associated with a feeling that you are not in full control.)

⎕ No discomfort at all
⎕ Minor discomfort
⎕ Mild discomfort
⎕ Moderate discomfort
⎕ Moderately severe discomfort
⎕ Severe discomfort
⎕ Very severe discomfort
15. When going to the toilet during the past week, have you had the SENSATION OF NOT COMPLETELY EMPTYING THE BOWELS? (This feeling of incomplete emptying means that you still feel a need to pass more stool despite having exerted yourself to do so.)

⎕ No discomfort at all
⎕ Minor discomfort
⎕ Mild discomfort
⎕ Moderate discomfort
⎕ Moderately severe discomfort
⎕ Severe discomfort
⎕ Very severe discomfort

